# Serum lipid profile changes and their clinical diagnostic significance in COVID-19 Mexican Patients

**DOI:** 10.1101/2020.08.24.20169789

**Authors:** Juan Fidel Osuna-Ramos, Horacio Rendón-Aguilar, Luis Adrián De Jesús-González, José Manuel Reyes-Ruiz, Arely Montserrat Espinoza-Ortega, Luis Antonio Ochoa-Ramírez, Alejandra Romero-Utrilla, Efrén Ríos-Burgueño, Alejandro Soto-Almaral, Juan José Ríos-Tostado, José Geovanni Romero-Quintana, Héctor Ponce-Ramos, Carlos Noe Farfan-Morales, Rosa María del Ángel, Héctor Barajas-Martínez, José Rodríguez-Millán, Jesús Salvador Velarde-Félix

## Abstract

**Background:** COVID-19 has been recognized as an emerging and rapidly evolving health condition. For this reason, efforts to determine changes in laboratory parameters of COVID-19 patients as biomarkers are urgent. Lipids are essential components of the human body, and their modulation has been observed implicated in some viral infections.

**Methods:** To evaluate the clinical diagnosis utility of the lipid profile changes in Mexican COVID-19 patients, the lipid profile of one hundred two COVID-19 positive patients from three hospitals in Culiacan, Sinaloa in northwest Mexico, was analyzed. ROC curves and binary logistic regression analysis were used as a predictive model to determine their clinical diagnostic utility.

**Results:** Significant changes in the serum lipid profile of patients with COVID-19, such as low levels of cholesterol, LDL, and HDL, while high triglycerides and VLDL were observed. The same abnormalities in the lipid profile among non-critical and critical COVID-19 patients were detected. The predictive model analysis suggests that cholesterol and LDL have AUC values of 0.710 and 0.769, respectively, for COVID-19 (p= 0.0002 and p= <0.0001), and LDL low levels might be a risk factor for critical COVID-19 (OR= 2.07, 95% IC: 1.18 to 3.63; p= 0.01).

**Conclusion:** Our findings suggest that low cholesterol and LDL levels could be considered an acceptable predictor for COVID-19, and low levels of LDL might be a risk factor for critical COVID-19 patients.

## 1. Introduction

Coronavirus disease 2019 (COVID-19) is a public health emergency of international concern caused by the severe acute respiratory syndrome coronavirus 2 (SARS-CoV-2). More than two hundred countries around the world have reported more than 13 million confirmed cases, and more than half a million deaths caused by COVID-19^1,2^. The epidemiological information of the COVID-19 pandemic in Latin America indicates that as of June nearly four million cases and more than two hundred thousand deaths have been reported^3^, while in Mexico, 311, 486 cases of COVID-19 and 36,327 deaths have been reported^4,5^. The SARS-CoV-2 transmission was initially linked to a local exotic animal market in the province of Wuhan, China, indicating a possible zoonosis ^6^ SARS-CoV-2 can be transmitted human-to-human, through the inhalation of aerosols with nasal discharge droplets, in addition to direct or indirect contact of the oral and nasal mucous membranes and eye membranes with everyday contaminated objects ^7-10^ The symptoms of the disease may be mild (flu-like infection with fever, headache or dry cough) or severe due to acute lung injury (ALI) and acute respiratory distress syndrome (ARDS), leading to acute breathing difficulties, respiratory failure, multiple organ failure, or death of the patient ^9-11^. Reports of patients with associated morbidity, mainly diabetes (DM), hypertension (HT), and other cardiovascular and cerebrovascular diseases, have been linked to poor prognosis of the disease^12-14^.

Additionally, manifestations of hypolipidemia were reported in Chinese patients with mild symptoms and progressively worsened according to the severity of the disease^15,16^. The severity of the disease could be attributed to alterations in cholesterol levels since mainly two groups of the population are more affected in morbidity and mortality by COVID-19^17-21^. On the other hand, changes in serum lipid profiles have been reported previously in other viral infections^22-26^, including respiratory viral infections, where their potential as metabolic biomarkers to help clinicians distinguish bacterial from viral infection in febrile patients has been determined^27^. For these reasons, we found it interesting to explore whether alterations in serum lipid levels in COVID-19 Mexican patients can be useful as biomarkers of disease or severity. In this study, we analyzed the lipid profile of one hundred two COVID-19 positive patients from 3 hospitals in Culiacan, Sinaloa, in northwest Mexico, one of the states with the highest number of cases of the disease in Mexico, and using a combination of ROC curves and logistic regression analysis we evaluate the clinical diagnosis utility of the lipid profile in Mexican COVID-19 patients.

## 1. Material and methods

### 2.1 Study design

A prospective and descriptive study was conducted at three different hospitals from Culiacan, Sinaloa, in northwest Mexico, from April 16 to June 16, 2020. All patients included in this study met the diagnostic criteria according to national and international guidelines for the diagnosis and treatment of COVID-19 ^27,28^, and the SARS CoV-2 infection was confirmed by the real-time polymerase chain reaction (RT-PCR) test. The information for all patients was obtained from electronic medical record systems, except the lipid profile (total serum cholesterol, low-density lipoprotein, high-density lipoprotein, triglycerides, and VLDL).

The risk stratification was determined according to the symptoms and clinical signs presented by the patients. In the mild disease with mild COVID-19, patients suffered of fever, headache, cough, respiratory symptoms, or chest pain, while in severe cases, patients have a respiratory rate (RR) ≥ 30 per minute or/and oxygen saturation ≤ 93 %. Critical COVID-19 patients met severe case criteria plus the need for mechanical ventilation, or/and had a shock or multiorgan failure^29^. Additionally, to dichotomize the variables and calculate risks and diagnostic accuracy, patients were divided into non-critical COVID-19 patients and critical COVID-19 patients: non-critical COVID-19 patients were those patients classified as mild or severe COVID-19 cases.

### 2.2 Serum Lipid Profile quantification

A fasting blood sample was taken from each participant at hospital admission to determine the total serum cholesterol (n= 102), low-density lipoprotein (LDL) (n= 94), high-density lipoprotein (HDL) (n= 93), and triglycerides (n= 98) levels by enzymatic method. Very-low-density lipoprotein (VLDL) (n= 98) levels were measured using Friedewald’s methodology ^30^

Additionally, the lipid profile data of a group of thirty healthy blood donors, who were part of a previous pre-pandemic study [32], was used as healthy controls. The control subjects were negative for a viral panel (anti-dengue IgM and IgG, HIV, HCV antibodies) and belonged to the same geographic region. They had no previous history of dyslipidemia or a history of cholesterol-lowering therapy.

The Research Ethical Committee of Hospital General de Culiacan "Bernardo J Gastelum" approved the study, and some patients writing informed consent, and in another, the consent was verbal.

### 2.3 Statistic analysis

The data were expressed as median and interquartile ranges (IQR, 25-75th percentile) to describe age, and mean and standard deviation (SD) were used to describe the lipid profile. The parametric Unpaired t-test with Welch’s correction was used to compare the lipid profile means of total COVID-19 patients between healthy controls, and the non-parametric U Mann Whitney test was used to compare age and lipid profile median values among non-critical and critical COVID-19 patients, using the GraphPad Prism software version 7. To determine if changes in the serum lipid profile are useful to predict COVID-19 disease or severity, receiver operating characteristic (ROC) curves and area under the curves ROC (AUC) were calculated to evaluate the sensitivity, specificity, positive (+LR) and negative (-LR) likelihood ratios to each lipid in the lipid profile in predicting COVID-19 disease and critical COVID-19. Moreover, binary logistic regression was used to calculate odds ratios (OR) and 95% confidence intervals (95% IC) between non-critical and critical COVID-19 patients based on the calculated optimal cut-off points using the MedCalc software version 14. Statistical significance was set at *p* < 0.05.

## 3. Results

During the study period, clinical and lipid profile information was obtained from 102 COVID-19 patients. Their demographic data showed a median age of 57 years and a higher frequency of male patients at hospital admission (Table 1). The patient stratification was according to the risk, based on the clinic, as shown in Table 1. Age differs significantly between non-critical and critical COVID-19 cases (p= 0.0095), and males were more frequent among non-critical and critical COVID-19 patients. About comorbidities, hypertension (HT), obesity, and diabetes (DM) were the most frequent, especially in the critical COVID-19 group of patients (Table 1).

**Table 1.** Demographic and clinical characteristics of COVID-19 patients. RR= respiration rate, SatO2= Oxygen Saturation, HT= Hypertension, DM= Diabetes.

|  | All patients<br>(n = 102) | Non-critical<br>COVID-19<br>(n = 64) | Critical COVID-19<br>(n = 38) |
| --- | --- | --- | --- |
| Age, median years (IQR) | 57 (45.75 – 64) | 54 (41.25 – 62) | 60.5 (51 – 69) |
| Sex, number of patients (%) |  |  |  |
| Female | 37 (36.3) | 24 (37.5) | 13 (34.2) |
| Male | 65 (63.7) | 40 (62.5) | 25 (65.8) |
| Clinical features, number (%) |  |  |  |
| RR ≥ 30 | 28 (27.5) | 12 (18.8) | 16 (42.1) |
| SatO<sub>2</sub> ≤ 93 % | 65 (63.7) | 37 (57.8) | 28 (73.7) |
| Intubation | 33 (86.8) | 0 | 33 (86.8) |
| Shock | 17 (16.7) | 0 | 17 (44.7) |
| Comorbidities, number (%) |  |  |  |
| HT | 59 (57.8) | 31 (48.4) | 28 (73.7) |
| DM | 37 (36.3) | 21 (32.8) | 16 (42.1) |
| Obesity | 44 (43.1) | 28 (43.8) | 16 (42.1) |

Regarding to the lipid profile in COVID-19 patients, the mean cholesterol (126 ± 41.02 mg/dL), LDL (69.5 ± 34.9 mg/dL) and HDL (29.9 ± 12.19 mg/dL) levels were lower compared to healthy control group (157.7 ± 37.1 mg/dL, LDL: 100.6 ± 27.64mg/dL and 41.43 ± 15.01 mg/dL, respectively) (Figure 1). On the other hand, both mean triglyceride (155.4 ± 72.49 mg/dL) and VLDL (31.09 ± 14.5 mg/dL) levels were higher in COVID-19 patients compared to healthy control (115 ± 65.07 mg/dL and 23 ± 13.01 mg/dL) (Figure 1).

**Figure 1.**
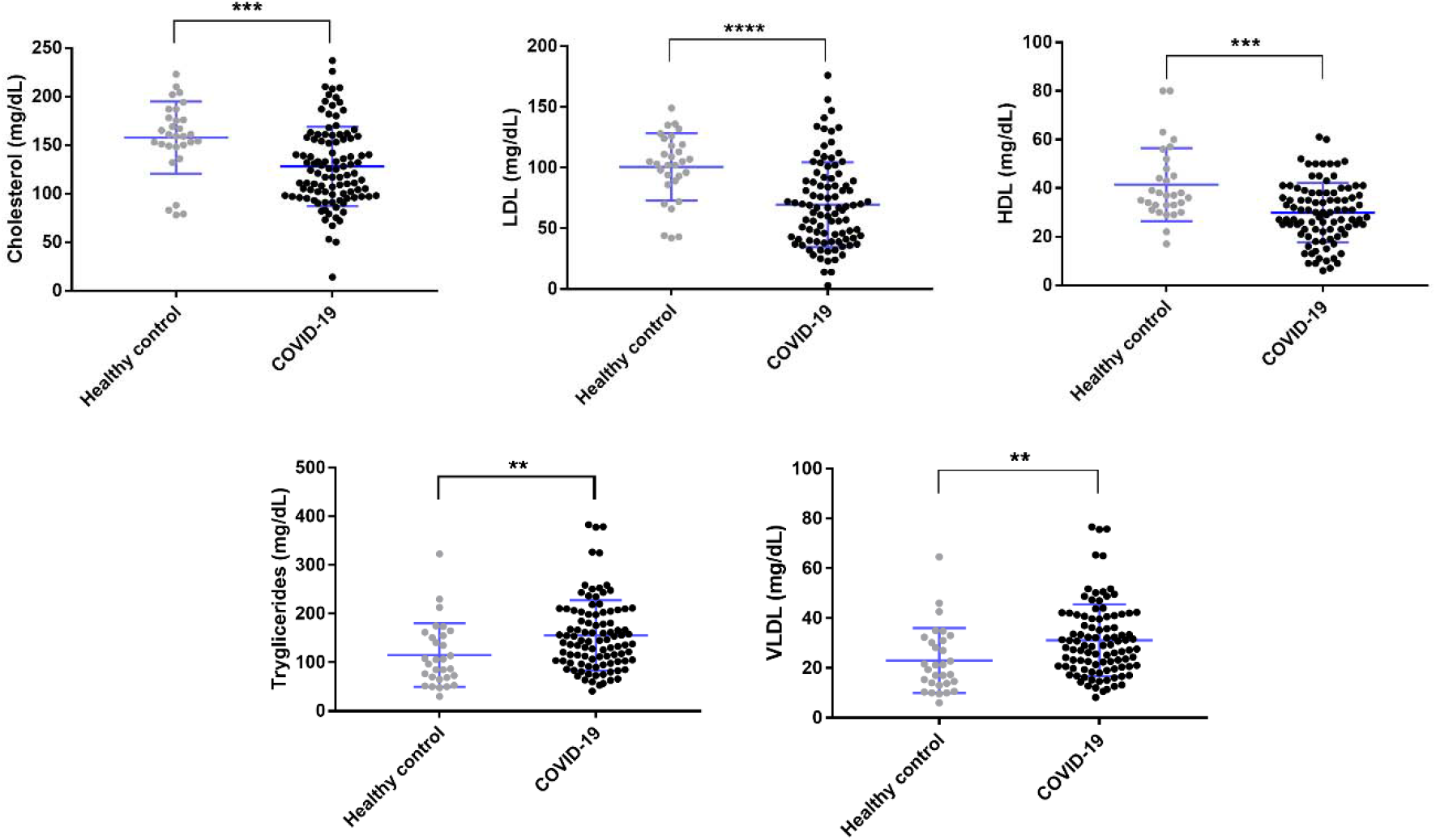
Lipid profile in COVID-19 patients (black circles) compared with healthy controls (Gray circles). In blue is showed the mean line and SD error bars lines. The output analysis is represented as the following p-values means differences: 0.0332 (*), 0.0021 (**), 0.0002 (***), <0.0001 (****).

To establish if differences between the lipid profile can accurately use to distinguish between COVID-19 patients and healthy individuals, ROC curves were used to determine optimal cut-off points and AUC for each lipid parameter. Based on Youden’s J index, the results of the ROC analysis, optimal cut-off, and the AUC calculations are shown in Table 2. Cholesterol and LDL showed optimal predictive value as well as higher sensitivity (greater than 60%) and specificity (greater than 80%) and AUC values of 0.710 and 0.769, respectively (Table 2).

**Table 2.** Optimal Cut-off point and AUCs predictors for the lipid profile in COVID-19 patients. +LR= positive likelihood ratio, -LR= negative likelihood ratio, AUC= area under the curve.

|  | Cut-off point | Sensitivity (95% CI) | Specificity (95% CI) | +LR (95% CI) | -LR (95% CI) | Youden Index J | AUC (95% CI) | p-value |
| --- | --- | --- | --- | --- | --- | --- | --- | --- |
| Cholesterol | ≤142 | 69.61 (59.7 - 78.3) | 80 (61.4 - 92.3) | 3.48 (1.7 - 7.2) | 0.38 (0.3 - 0.5) | 0.4961 | 0.710 (0.625 - 0.786) | 0.0002 |
| LDL | ≤85 | 72.34 (62.2 - 81.1) | 80 (61.4 - 92.3) | 3.62 (1.7 - 7.5) | 0.35 (0.2 - 0.5) | 0.5234 | 0.769 (0.684 - 0.840) | <0.0001 |
| HDL | ≤28 | 48.39 (37.9 - 59.0) | 93.33 (77.9 - 99.2) | 7.26 (1.9 - 28.1) | 0.55 (0.4 - 0.7) | 0.4172 | 0.719 (0.630 - 0.796) | <0.0001 |
| Triglycerides | >114 | 67.35 (57.1 - 76.5) | 63.33 (43.9 - 80.1) | 1.84 (1.1 - 3.0) | 0.52 (0.3 - 0.8) | 0.3068 | 0.679 (0.591 to 0.759) | 0.0023 |
| VLDL | >22.8 | 67.35 (57.1 - 76.5) | 63.33 (43.9 - 80.1) | 1.84 (1.1 - 3.0) | 0.52 (0.3 - 0.8) | 0.3068 | 0.679 (0.591 to 0.759) | 0.0023 |

In further analysis, to determine if there were differences between the lipid profile in COVID-19 patients according to severity, non-critical and critical. Lower cholesterol (119 mg/dL, IQR: 94.5 - 154.5 mg/dL, n= 38), LDL (50 mg/dL, IQR: 33 - 87.75 mg/dL, n= 36), and HDL (26 mg/dL, IQR: 16-43 mg/dL, n= 35) values were observed in critical compared with non-critical patients (cholesterol: 125 mg/dL, IQR: 102 - 160.8, n= 64; LDL: 69 mg/dL, IQR: 48.25 - 95.25 mg/dL, n= 58; HDL: 30 mg/dL, IQR: 22 - 36.5 mg/dL, n= 61), while median values of triglycerides (145 mg/dL, IQR: 100.5 - 201.5 mg/dL, n= 37) and VLDL in critical patients were higher than median triglycerides and VLDL values in no-critical (138 mg/dL, IQR: 103 – 105 mg/dL, n= 61, and 27.6 mg/dL, IQR: 20.6 - 41 mg/dL, for triglycerides and VLDL, respectively). However, only median LDL values differences were statistically significant among non-critical and critical COVID-19 patients (Figure 2).

**Figure 2.**
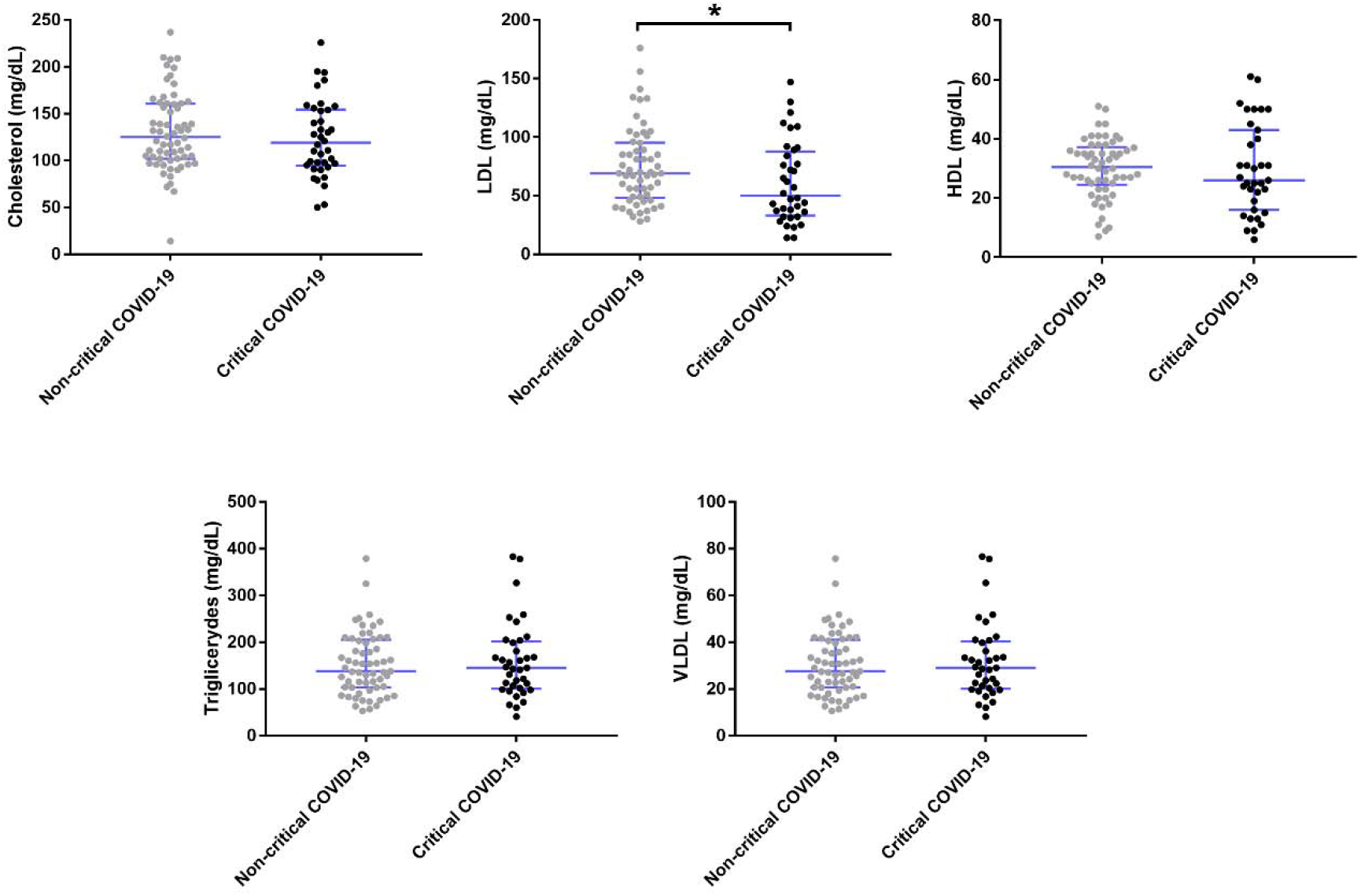
Lipid profile compared between no-critical (gray) and critical (black) COVID-19 patients. In blue is showed the median line and IQR error bars lines. The output analysis is represented as the following p-values mean differences: 0.0332 (*).

Moreover, a combination of ROC curves and binary logistic regression analysis was performed as a predictive model. The optimal working point was for LDL and showed predictive values of 50% sensitivity and 75% specificity, as well as an AUC of 0.621, with an optimal cut-off value ≤ 48 mg/dL, to discriminate between non-critical and critical COVID-19 (Table 3).

**Table 3.** Optimal Cut-off point and AUCs predictors for critical COVID-19 patients. +LR= positive likelihood ratio, -LR= negative likelihood ratio, AUC= area under the curve

|  | Cut-off point | Sensitivity (95% CI) | Specificity (95% CI) | +LR (95% %) | -LR (95% %) | Youden Index J | AUC (95% CI) | p-value |
| --- | --- | --- | --- | --- | --- | --- | --- | --- |

**Table 3.** Optimal Cut-off point and AUCs predictors for critical COVID-19 patients.
|  |  |  |  | CI) | CI) |  |  |  |
| --- | --- | --- | --- | --- | --- | --- | --- | --- |
| Cholesterol | ≤99 | 36.84<br>(21.8 -<br>54.0) | 78.12<br>(66.0 -<br>87.5) | 1.68<br>(0.9 -<br>3.1) | 0.81<br>(0.6 -<br>1.1) | 0.1497 | 0.557<br>(0.45<br>5 to<br>0.655<br>) | 0.344<br>4 |
| LDL | ≤48 | 50.00<br>(32.9 -<br>67.1) | 75.86<br>(62.8 -<br>86.1) | 2.07<br>(1.2 -<br>3.6g<br>) | 0.66<br>(0.5 -<br>0.9) | 0.2586 | 0.621<br>(0.51<br>5 to<br>0.719<br>) | 0.054<br>2 |
| HDL | ≤25 | 48.57<br>(31.4 -<br>66.0) | 71.19<br>(57.9 -<br>82.2) | 1.69<br>(1.0 -<br>2.9) | 0.72<br>(0.5 -<br>1.0) | 0.1976 | 0.536<br>(0.43<br>0 to<br>0.639 | 0.599<br>6 |
| Triglyceride<br>s | >251 | 13.51<br>(4.5 -<br>28.8) | 95.08<br>(86.3 -<br>99.0) | 2.75<br>(0.7 -<br>10.8<br>) | 0.91<br>(0.8 -<br>1.0) | 0.0859<br>5 | 0.511<br>(0.40<br>8 to<br>0.613<br>) | 0.856<br>8 |
| VLDL | >50.<br>2 | 13.51<br>(4.5 -<br>28.8) | 95.08<br>(86.3 -<br>99.0) | 2.75<br>(0.7 -<br>10.8<br>) | 0.91<br>(0.8 -<br>1.0) | 0.0859<br>5 | 0.511<br>(0.40<br>8 to<br>0.613<br>) | 0.856<br>8 |

On the other hand, the binary logistic regression analysis on the correlations showed that LDL ≤ 48 mg/dL cut-off value might be a risk factor for critical COVID-19 (OR= 2.07, 95% IC: 1.18 to 3.63; p= 0.01) (Table 4).

**Table 4.** Binary logistic regression analysis of lipid profile associated with critical COVID-19 based on the optimal cut-off values. OR= odd ratio, 95% IC =95% Interval confidence, SE= standard error.

| Variable, cut-off point | OR | 95% IC | P value |
| --- | --- | --- | --- |
| Cholesterol, ≤99 | 1.68 | 0.90 - 3.13 | 0.10 |
| LDL, ≤48 | 2.07 | 1.18 - 3.63 | 0.01 |
| HDL, ≤25 | 1.76 | 1.02 - 3.01 | 0.04 |
| Triglycerides, >251 | 2.74 | 0.69 - 10.83 | 0.13 |
| VLDL, >50.2 | 2.74 | 0.69 - 10.83 | 0.13 |

## 4. Discussion

Lipids are essential components of the human body, and they are involved in metabolic diseases such as diabetes, which is characterized by elevated total cholesterol, LDL, HDL, and triglycerides serum levels ^31,32^. Also, increased serum levels of cholesterol, triglycerides, LDL and decreased HDL are known to be associated with the significant risk factors for Cardiovascular Disease and hypertension^33^. Interestingly, changes in serum lipid profiles are prognostic indicators of diseases, including viral infections. In this sense, clinical research indicates that patients infected with human immunodeficiency (HIV)^22^, dengue (DENV) ^23-25^, and hepatitis B (HBV) ^26^ viruses develop hypocholesterolemia and hypertriglyceridemia, as well as low levels of LDL or HDL.

A recently published study performed in China found that LDL and total cholesterol levels were significantly lower in COVID-19 patients as compared with the control group ^15^. In the same way of evidence, Fan et al. reported a similar pattern of low cholesterol, LDL, and HDL levels in a group of 21 patients with COVID-19 in Wuhan, China ^16^ Therefore, it appears that there is a pattern of dyslipidemia that may be characteristic of patients with COVID-19. However, it is true that worldwide there is a wide variation in serum lipid profile levels depending on the different population groups^33^.

In the present study, conducted in a Mexican population, significant changes in the lipid profile of patients with COVID-19 was found. The same pattern of dyslipidemia observed in previously reported studies in Asia was also observed in the Mexican population ^15,16^, as low levels of cholesterol, LDL, and HDL, while high levels of triglycerides and VLDL when compared with a group of healthy individuals. This repeating pattern in lipid profile changes in COVID-19 patients made us propose the lipid profile changes as a possible biomarker for COVID-19. Therefore, through a predictive model using ROC curves, we were able to detect the optimal cut-off points of ≤99 mg/dL for cholesterol and ≤48 mg/dL for LDL, which could be considered acceptable predictors for COVID-19 since they demonstrated to have significant AUC values between 0.70 and 0.80 for diagnostic accuracy ^34^.

Serum dyslipidemia during the COVID-19 may be due to the role of lipids in SARS-Cov-2 replication in the host cell since they participate in the internalization of the virus into the cell^35–37^, and in the dynamism of the lipid rafts in the membrane where the ACE-2 receptor is located ^38,39^. Also, SARS-CoV-2 regulates the lipidic metabolism to generate viral membrane and pack its genome and in the exocytosis of the virions ^40,41^. For this reason, a growing recent interest in the lipids changes research has been noted in SARS-CoV-2 infected patients, since people with metabolic diseases such as DM, HT, and obesity, possibly leading to a higher risk of complications and mortality from COVID-19 ^42,43^ Our study found high frequencies of comorbidities alone or in combination, such as HT, DM, and obesity in COVID-19 patients. One of the main concerns of the Institute of Public Health is the high prevalence of these comorbidities in the Mexican population ^44,45^.

At this moment, COVID-19 is considering as an emerging and rapidly evolving condition. So, efforts to determine the change in laboratory parameters as biomarkers have been directed towards the identification of severe or critical cases of COVID-19 ^46,47^ In this regard, we stratified into non-critical and critical patients, and the same pattern of low levels of cholesterol, LDL, and HDL was observed in critical COVID-19 patients.

The above is in concordance with observed by Wei et al. previously reported^15^. They divided COVID-19 patients per their severity into mild, severe, and critical categories. They found that the development of hypolipidemia begins in patients with mild symptoms and progressively worsens in association with the severity of the disease ^15^ However, they reported that triglyceride levels decreased significantly in critical cases compared to mild and severe cases. Contrary to our results, as higher triglyceride levels were found in critically ill patients with COVID-19, but only median LDL levels differences were statistically different between non-critical and critical patients. In another similar research, Fan et al., besides reporting a pattern of dyslipidemia similar to what has previously been observed in patients with COVID-19, they also showed that degrees of decrease in LDL levels are highly likely to be associated with the severity and mortality of the disease^16^. The optimal cut-off point was for LDL in the ROC analysis, and we observed that LDL might be a risk factor for critical COVID-19. Because of the low AUC value obtained for LDL, it could not be considered an acceptable biomarker to differentiate between non-critical and critical COVID-19 patients. This may be due to the limitations of our study, such as sample size or the low statistical power difference observed between both groups of patients. However, it cannot be ruled out that LDL levels may be useful for decision making in the evolution of COVID-19 critically ill patients, and more studies are needed, especially in high-risk populations.

## Data Availability

No suplementary material is included

## Acknowledgments

Juan Fidel Osuna-Ramos, Luis Adrián De Jesús-González, José Manuel Reyes-Ruiz and Carlos Noe Farfan-Morales received scholarships from CONACYT.

